# Neuroinflammation predicts disease progression in progressive supranuclear palsy

**DOI:** 10.1101/2020.05.19.20106393

**Authors:** Maura Malpetti, Luca Passamonti, P. Simon Jones, Duncan Street, Timothy Rittman, Tim D. Fryer, Young T. Hong, Patricia Vázquez Rodríguez, W. Richard Bevan-Jones, Franklin I. Aigbirhio, John T. O’Brien, James B. Rowe

## Abstract

**Objective:** In addition to tau pathology and neuronal loss, neuroinflammation occurs in progressive supranuclear palsy (PSP). We test the hypotheses that baseline *in vivo* assessments of regional neuroinflammation ([^11^C]PK11195 PET), tau pathology ([^18^F]AV-1451 PET), and atrophy (structural MRI) predict disease progression.

**Methods:** Seventeen patients with PSP-Richardson’s syndrome underwent a baseline multi-modal imaging assessment. Disease severity was measured at baseline and serially up to 4 years with the PSP-rating-scale (average interval 5 months). Regional grey-matter volumes and PET ligand binding potentials were summarised by three Principal Component Analyses (PCAs). A linear mixed effects model was applied to the longitudinal PSP-rating-scale scores. Single-modality imaging predictors were regressed against the individuals’ estimated rate of progression to identify the prognostic value of baseline imaging markers.

**Results:** The PCA factors reflecting neuroinflammation and tau burden in the brainstem and cerebellum correlated with the subsequent annual rate of change in the PSP-rating-scale. PCA-derived PET markers of neuroinflammation and tau pathology correlated with brain atrophy in the same regions. However, MRI markers of brain atrophy alone did not predict clinical progression.

**Conclusions:** Molecular imaging with PET can predict clinical progression in PSP. These data encourage the evaluation of immunomodulatory approaches to disease-modifying therapies in PSP, and the potential for PET to stratify patients for early phase clinical trials.

## Introduction

Neuroinflammation has been recognized as an common pathogenic process in Progressive Supranuclear Palsy (PSP) and other tauopathies such as Alzheimer’s disease^1^, with genetic, epidemiological, and imaging associations. For example, activated microglia are found in the neighbourhood of neurofibrillary tangles, even during early stages of disease^2,3^, and are directly synaptotoxic^1^. Neuroinflammation, including microglial activation, interacts with tau pathology to promote cell dysfunction and death in preclinical models of tauopathy.

PET radioligands have been developed to assess neuroinflammation and tau pathology accumulation *in vivo* in clinical cohorts. [^11^C]PK11195 is a widely used PET tracer, that binds primarily to activated microglia in PSP^4–6^ and other neurodegenerative disorders^7^. The ligand [^18^F]AV-1451 is widely to assess tau pathology in Alzheimer’s disease, and can be informative about tau pathology in PSP^6,8–14^ despite lower sensitivity to the tau isoforms in PSP, and off target binding in some regions^15,16^. However, it has not been shown whether either of these PET biomarkers of neuroinflammation and tau pathology predict longitudinal clinical progression in these patients.

Our main hypothesis was that inflammation in the subcortical regions associated with PSP pathology promote disease progression. We therefore test whether baseline *in vivo* measures of neuroinflammation ([^11^C]PK11195 PET) predict the annual rate of clinical progression in patients with PSP-Richardson’s syndrome. We test secondary hypotheses regarding the predictive value of baseline inflammation elsewhere, tau pathology ([^18^F]AV-1451 PET) and atrophy (structural MRI).

## Methods

### Participants

As part of the Neuroimaging of Inflammation in Memory and Other Disorders (NIMROD) study^17^, we recruited 17 people with a clinical diagnosis of probable PSP according to Movement Disorder Society (MDS) 1996 criteria^18^. These participants also met the later MDS-PSP 2017 criteria for PSP-Richardson’s syndrome^19^. The cross-sectional baseline data have been published previously^6^. Participants underwent a baseline neuropsychological assessment, followed by a MRI scan and two PET scans with [^11^C]PK11195 and [^18^F]AV-1451, to respectively assess neuroinflammation and tau pathology. Disease severity was measured at the baseline visit an serially up to 4 years using the PSP rating scale (PSP-RS)^20^. Assessments were at an average of 5-months intervals (standard deviation (SD) ± 2.3 months). *Post mortem* confirmation of PSP pathology was available in 8 patients, and for all 17 participants the clinical diagnosis was reviewed and confirmed at follow-up.

Participants had mental capacity and provided written informed consent. The NIMROD protocol was approved by the National Research Ethic Service’s East of England Cambridge Central Committee and the UK Administration of Radioactive Substances Advisory Committee.

### MRI and PET data acquisition and pre-processing

Full details of the imaging protocols have been published elsewhere^5,8^. In brief, patients underwent 3T MRI, together with dynamic PET imaging of [^11^C]PK11195 and [^18^F]AV-1451 for 75 and 90 minutes, respectively. MP-RAGE T1-weighted MRI was acquired on Siemens Magnetom Tim Trio and Verio scanners (Siemens Healthineers, Erlangen, Germany), while PET scans were performed on a GE Advance and a GE Discovery 690 PET/CT (GE Healthcare, Waukesha, USA). The use of identical emission data acquisition protocols and image reconstruction algorithms on the two PET scanners meant that the differences were effectively limited to the attenuation correction method (rotating rod ^68^Ge/^68^Ga transmission scan vs. a low dose CT scan) and the axial spatial resolution (6.8 mm FWHM vs. 5.1 mm FWHM). Regarding the attenuation correction, the CT (Hounsfield unit) to 511 keV linear attenuation coefficient transformation used on GE PET/CT systems is that of Burger et al^21^, which was determined from data acquired with a GE Discovery LS PET/CT, the PET part of which is identical to the GE Advance, thereby enhancing the correspondence between GE PET and PET/CT systems. With respect to differences in spatial resolution, the primary data given in the paper are for large regions of interest. This will limit the impact of any spatial resolution differences, which for brain imaging on the scanners used mainly occur in the axial dimension. Furthermore, patient motion, together with resolution losses in image processing steps, such as realignment of dynamic image series and co-registration to MR, will reduce these differences. Median (mean and standard deviation) of the time interval between the baseline clinical assessment and the imaging scans were: 0.0 (1.1 ± 1.5) months for MRI, 2.0 (2.7±2.0) months for [^11^C]PK11195 PET, and 1.0 (1.9±1.8) months for [^18^F]AV-1451 PET.

For each subject, the aligned dynamic PET image series for each scan was rigidly co-registered to the T1-weighted MRI image. Grey matter volumes and non-displaceable binding potential (BP_ND_) values for each tracer were calculated in 83 cortical and subcortical ROIs using a modified version of the Hammersmith atlas (www.brain-development.org), which includes parcellation of the brainstem and cerebellar dentate nucleus. Each T1 image was spatially normalised using ANTS (http://www.picsl.upenn.edu/ANTS/) and the inverse transform was applied to a version of the Hammersmith atlas to bring the regions of interest to native T1 space. The T1-weighted images were segmented into grey matter, white matter and cerebrospinal fluid (CSF) with SPM12 (www.fil.ion.ucl.ac.uk) and used to determine regional grey matter, white matter and CSF volumes, and to calculate the total intracranial volume (TIV = grey matter + white matter + CSF) in each participant. Prior to kinetic modelling, regional PET data were corrected for partial volume effects from cerebrospinal fluid by dividing by the mean regional grey-matter plus white-matter fraction determined from SPM segments smoothed to PET spatial resolution. For [^11^C]PK11195, supervised cluster analysis was used to determine the reference tissue time-activity curve and BP_ND_ values were calculated in each ROI using a simplified reference tissue model with vascular binding correction^22^. For [^18^F]AV-1451, BP_ND_ values were quantified in each ROI using a basis function implementation of the simplified reference tissue model^23^, with superior cerebellar cortex grey matter as the reference region. This cerebellar region was selected as reference region given *post mortem* evidence showing minimal tau pathology in PSP (see pathology data in Supplementary material in Passamonti et al.^8^).

### Statistical analyses

Grey matter volumes and BP_ND_ values for each ligand were combined across the two hemispheres to derive 43 bilateral whole-brain regions of interest (ROIs)^5,6,8^, which were next included in separate PCAs for each imaging modality. Varimax rotation was applied in all PCAs to maximize interpretability and specificity of the resulting components. The components with eigenvalues > 1 were retained, explaining >80% of the cumulative variance.

A linear mixed model was applied to the longitudinal PSP-RS scores collected from the first research visit to estimate the clinical annual rate of change at group level, and then extract a patient-specific estimate of disease progression. The model included the estimation of a random intercept and slope, with time (in years) as independent variable and PSP-RS scores as dependent variable (formula: PSP-RS ~ Time + (Time | ID)). The effect of time on clinical changes has been also tested via likelihood ratio tests of the model described above against the null model without the time effect. The linear mixed effects analysis was performed using R 3.6.3 and lme4 package (R Core Team, 2012).

To test whether specific neuroanatomical patterns of grey matter atrophy, microglial activation and tau pathology predict clinical progression, linear regression models were applied with the estimated rate of change (slope) as dependent variable, and each method specific PCA component as predictor. First, we tested for significant univariable regressions on slope with each modality-specific subcortical component as predictor, accordingly with our main hypothesis. Then, we explored the predictive value of cortical components running separate linear regression analyses for each imaging method and component. Age, education and sex were included as covariates of no interest. We tested the correlations between rate of change of clinical scores, the disease duration and the first raw PSP-RS score at the baseline research visit.

Analogous linear regression models were the estimated with the intercept of the clinical severity as dependent variable. This identifies a cross-sectional association between imaging markers and clinical severity at baseline, which was robustly estimated at individual level from the linear mixed effects model on longitudinal PSP-RS scores. For cross-sectional analyses, we expected to find significant associations with subcortical imaging components^10,11^.

Lastly, the modality-specific subcortical components were included in cross-modality Pearson correlations to test for associations between the strength of regional atrophy, neuroinflammation and tau pathology.

### Data availability statement

Anonymized data may be shared through the Dementias Platform UK Portal by request to the senior author from a qualified investigator for non-commercial use (data sharing is subject to participants’ consent and GDPR regulations).

## Results

The demographics, clinical and cognitive variables of our sample are summarized in Table 1. Fifteen out of 17 patients died within 5 years from the baseline assessment (median = 2.4 years; mean ± SD [range] = 2.2 ± 1.2 [0 – 4.5] years from baseline visit). In Table 1, we report demographic, clinical characteristics and group comparisons for two subgroups of patients, identified dividing the total group on the median of the time interval between study baseline and death in years. Age, years of education, baseline PSP-RS, and annual rate of change in PSP-RS were compared with independent-samples t-tests; sex was compared with the Chi-square test.

**Table 1.** Demographic and clinical characteristics for the total patient group, and for sub-groups split based on patient survival from study baseline relative to the median time interval between study baseline and death (median = 2.4 years; mean ± standard deviation = 2.2 ± 1.2).

|  | **Total patient group** | **Patient survival < 2.4 years** | **Patient survival > 2.4 years** | **Difference** |
| --- | --- | --- | --- | --- |
| **N** | 17 | 8 | 9 |  |
| **Sex**<br>(F/M) | 7/10 | 3/5 | 4/5 | $\chi^2=0.08$, $p=0.772$ |
| **Age**<br>(mean ± SD) | 68.3±5.7 | 68.8±7.5 | 67.9±3.8 | $t(15)=0.30$, $p=0.776$ |
| **Education**<br>(mean ± SD) | 12.1±1.9 | 11.9±1.9 | 12.3±2.0 | $t(15)=-0.48$, $p=0.635$ |
| **Disease Duration**<br>(mean ± SD) | 4.7±1.8 | 5.0±1.8 | 4.5±1.8 | $t(15)=-0.57$, $p=0.577$ |
| **PSP-RS baseline**<br>(mean ± SD) | 41.2±14.5 | 46.0±15.6 | 37.0±12.8 | $t(15)=1.31$, $p=0.211$ |
| **Clinical progression – PSP-RS points/year**<br>(mean ± SD) | 6.2±1.5 | 6.4±0.8 | 5.9±1.9 | $t(15)= 0.73$, $p=0.478$ |

Abbreviations: PSP-RS=Progressive Supranuclear Palsy – Rating Scale; SD=standard deviation; $t()$ = $t$ -test; $p$ = $p$ -value

### Principal component analysis of grey matter volumes, [^11^C]PK11195 BP_ND_ and [^18^F]AV-1451 BP_ND_

For grey matter volumes, seven components were identified, which explained 80.3% of the total data variance. Figure 1 (left panel) provides a pictorial representation of the first four components and Supplementary Material (SM) Table 1 gives details on regional weights in all seven components. Component 1 was widely distributed, including medial frontal cortex, and thalamus, occipito-parietal regions, posterior cingulate cortex, and post-central cortex, (32.0% of the total variance). Component 2 (11.8% variance) was weighted to the midbrain, substantia nigra and pons in the brainstem, nucleus accumbens and putamen in the striatum as well as to the amygdala, hippocampus and pre-central cortex, cerebellar grey-matter and dentate gyrus. Component 3 (10.0% variance) loaded onto orbitofrontal cortex, anterior temporal lobe and lingual gyrus. Component 4 (7.9% variance) encompassed the superior temporal gyrus, fusiform gyrus, middle inferior temporal lobe, and insula.

**Figure 1.**
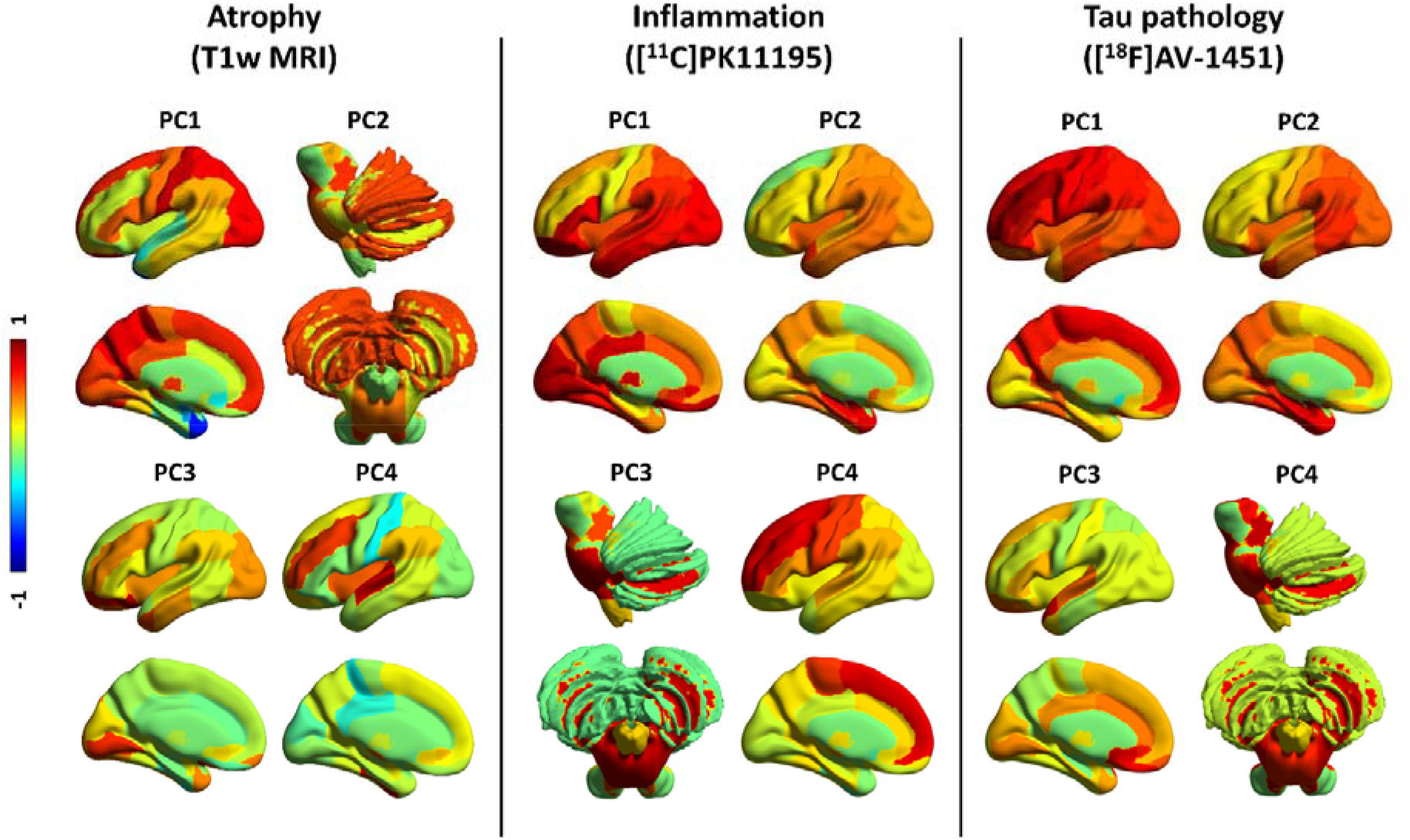
First four principal components (PC) for grey matter volumes (left panel), [^11^C] PK11195 non-displaceable binding potential (BP_ND_ – middle panel), and [^18^F]AV-1451 BP_ND_ (right panel). The colours represent the rotated weights (range: from -1 to 1) of the brain regions for each component.

For [^11^C]PK11195 BP_ND_ and [^18^F]AV-1451 BP_ND_, each PCA identified four components, which collectively and respectively explained 81.4% and 81.8% of the data variance, as reported in Malpetti et al.^6^. For [^11^C]PK11195 (Figure 1, middle panel; SM Table 2), component 1 loaded onto posterior cortical regions, the orbitofrontal cortex and cerebellar grey-matter; component 2 grouped together medial and superior regions of the temporal lobe, insula and temporo-parietal junction; component 3 was weighted to brainstem regions (i.e., midbrain and pons), the dentate nucleus, and cerebellar white-matter; while component 4 included the superior and medial frontal regions. For [^18^F]AV-1451 (Figure 1, right panel; SM Table 3), component 1 reflected global cortical binding; component 2 grouped insula and medial temporal lobe regions; component 3 loaded onto the anterior superior temporal gyrus and frontal subgenual cortex; component 4 was weighted towards subcortical areas including the midbrain, pons, substantia nigra, thalamus, dentate nucleus, and cerebellar white matter.

### Linear mixed model on longitudinal PSP-RS

The linear mixed model on longitudinal PSP-RS scores, considering the first research visit as baseline score, indicated a significant effect of time (Mean=6.15 points/year, SD=1.06, Figure 2). The model comparison against the null model confirmed the significant effect of time (Δχ^2^ = 42.61, Δdf=3, p<0.0001). We applied an analogous model that included all PSP-RS scores available from patients’ initial clinical diagnosis visit to their latest clinical visit, confirming a similar annual rate of change in PSP-RS (Mean=7.20 points/year; SD=1.18).

**Figure 2.**
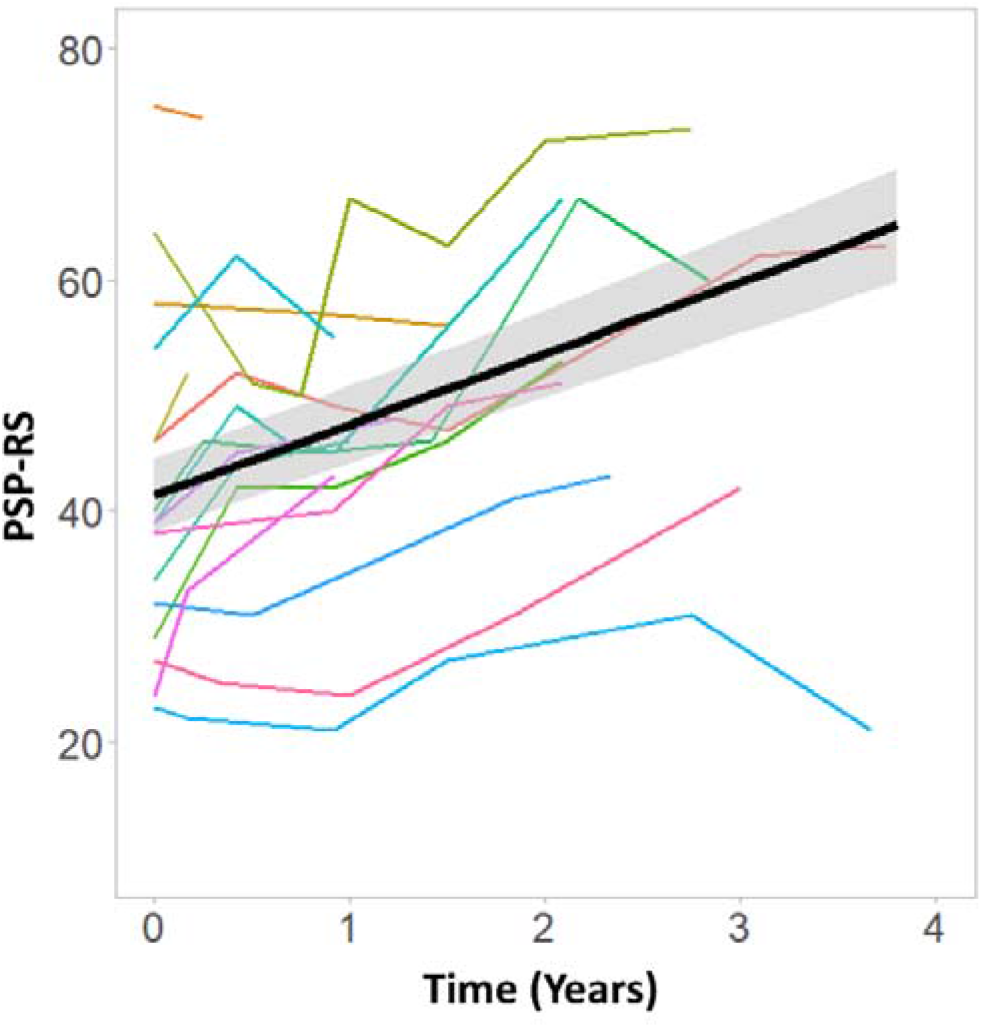
Clinical severity as measured by the Progressive Supranuclear Palsy Rating Scale (PSP-RS) over time. Coloured lines chart the time-course of PSP-RS score in individual patients. The black line represents the linear fit at group level, which shows an annual rate of change of 6.15 (p<0.0001) points in PSP-RS.

### Single-modality linear regressions

We tested whether imaging markers in subcortical components predicted longitudinal PSP-RS progression, applying univariable linear regression models for each of the three modality-specific subcortical components (MRI components #2, [^11^C]PK11195 component #3 and [^18^F]AV-1451 component #4). Correcting for age, education and sex, the annual rate of clinical progression was related positively with: 1) the [^11^C]PK11195 subcortical component #3 (Std Beta=0.624, p=0.023 and 2) the [^18^F]AV-1451 subcortical component #4 (Std Beta=0.840, p=0.003) (Figure 3, top row). Applying univariable regression models on slope with single cortical components, age, education and sex as predictors, no other components had significant correlations with the annual rate of clinical progression (p>0.05 after FDR correction for multiple comparisons). For MRI, no component showed an association with clinical rate of change (p>0.05 after FDR correction for multiple comparisons). The univariable regression models on clinical progression were not significant for disease duration (Std Beta=-0.08, p=0.775 uncorrected), or the baseline PSP-RS score (Std Beta=0.33, p=0.196) as single regressors.

**Figure 3.**
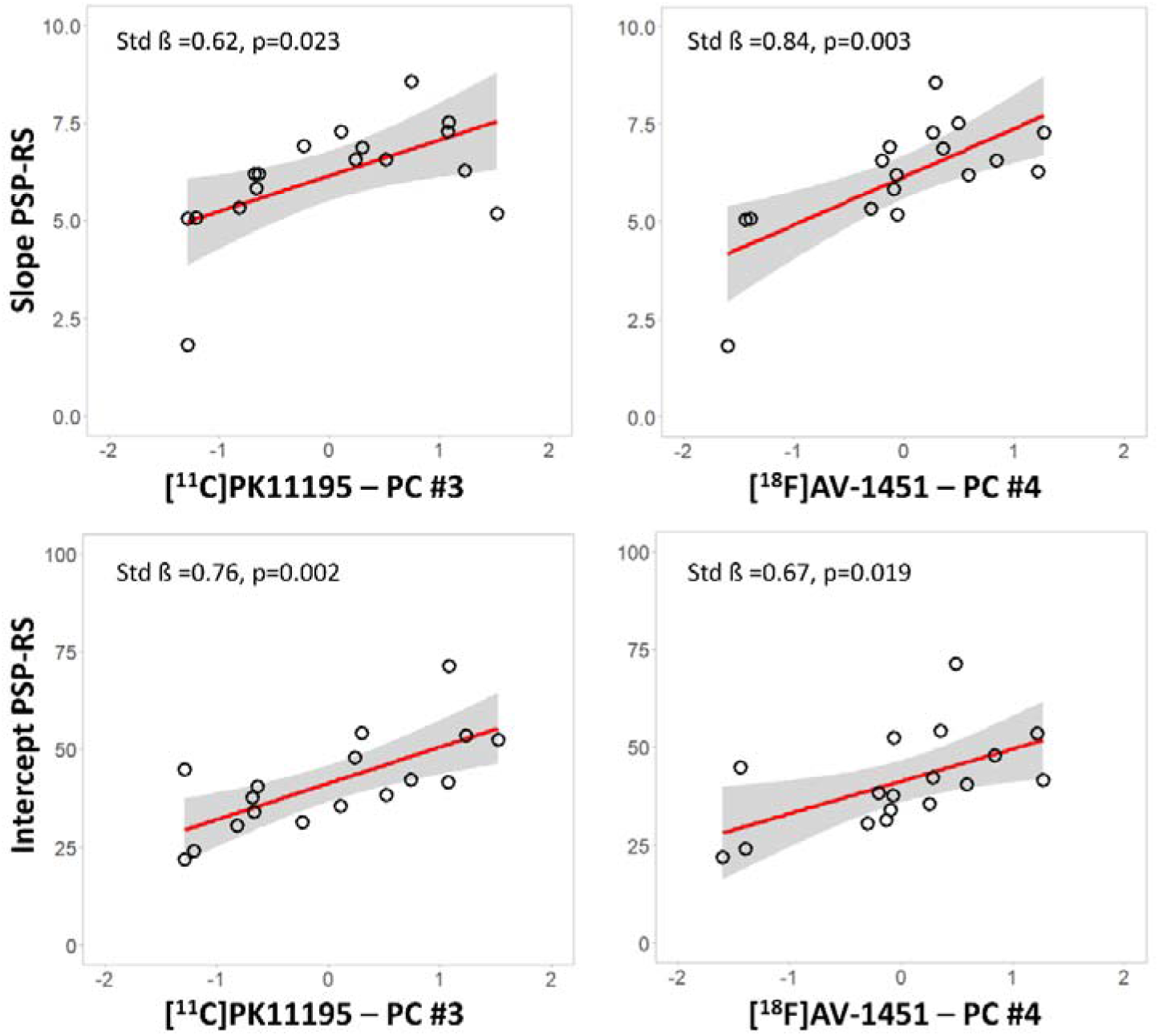
Significant regression analyses of annual change in Progressive Supranuclear Palsy Rating Scale (PSP-RS) scores (top row) and intercept PSP-RS scores (bottom row) against baseline scores for each modality-specific principal component (x axis – residual component values corrected for covariates): [^11^C]PK11195 PET (left panel), and [^18^F]AV-1451 PET (right panel). Estimated parameters are reported for each model with age, education and sex as covariates.

We tested whether imaging markers in subcortical components were related to baseline variation in disease severity. Across the three modality-specific subcortical components, univariable linear regression models with individual PSP-RS *intercept* scores as the dependent variable, correcting for age, education and sex, indicated significant associations with the [^11^C]PK11195 subcortical component #3 (Std Beta=0.755, p=0.002), and for [^18^F]AV-1451 subcortical component #4 (Std Beta=0.673, p=0.019) (Figure 3, bottom row). However, there was no significant association between intercept and any of the grey-matter components or cortical PET components (all p>0.05 after FDR correction for multiple comparisons). The univariable regression model on clinical intercept scores with disease duration (Std Beta=0.45, p=0.071) was not significant.

### Inter-modality correlations between subcortical imaging components

Simple correlations across subjects between MRI and PET subcortical components of clinical slope were significant for MRI component #2 with [^11^C]PK11195 component #3 (R=-0.584, p=0.014 uncorrected, p=0.014 FDR correction), and [^18^F]AV-1451 component #4 (R=-0.626, p=0.007 uncorrected, p=0.011 FDR correction). The correlation between [^11^C]PK11195 component #3 and [^18^F]AV-1451 component #4 was also significant (R=0.769, p<0.0001 uncorrected, p<0.0001 FDR correction)^6^.

## Discussion

The main finding of this study is that subcortical neuroinflammation is associated with clinical severity at baseline and faster clinical progression of PSP. A similar effect is seen for the estimated subcortical tau pathology, with the caveats related to interpreting [^18^F]AV1451 binding in PSP. The PET markers were associated with each other and with structural MRI measures for atrophy in the same regions. However, subcortical grey matter atrophy was not correlated with subsequent clinical progression and was not significantly related to clinical severity at the baseline. Similarly, clinical severity at baseline was not predictive of clinical progression in the following years, suggesting that the annual rate of clinical changes is approximately constant throughout different stages of disease.

Several studies have explored the association between changes in clinical severity and *in vivo* neuroimaging markers for microglial activation^4^, tau pathology^8–12^ and atrophy^24–27^. This study however focusses on the prognostic (or predictive) potential of baseline multimodal imaging markers. The baseline uptake of both [^11^C]PK11195 and [^18^F]AV-1451 in PSP-related subcortical regions were correlated with the subsequent annual rate of change in severity, as measured by the PSP-RS. Note that we are not testing whether the progression of PET markers compares with progression of disease severity. The progression of [^18^F]AV-1451 uptake has been compared with the progression of MRI measured of atrophy^13^, with greater changes in atrophy than changes in PET signals over time. But, for prognostic value, we found that grey matter volumetric measures were weaker predictors than the PET markers, despite the correlation of subcortical gray matter volumes with [^11^C]PK11195 and [^18^F]AV-1451 binding. The latter correlation suggests a close relationship between not only microglial activation and tau pathology^2,6^, but also with neurodegeneration in the subcortical regions most often associated with the pathological hallmarks of PSP^28^. Our findings align with studies of another tauopathy, Alzheimer’s disease, in which the baseline *in vivo* PET markers of tau pathology and microglial activation predicted clinical progression^29,30^, which also outperformed structural MRI predictors^30^.

The correlation of *in vivo* PET measures with baseline disease severity has been reported in previous studies^10,11^. Using [^11^C]PK11195 PET to assess neuroinflammation, a positive association was observed between clinical severity, assessed with PSP-RS, and ligand binding in the pallidum, midbrain and pons^5^. There was a strong association between [^11^C]PK11195 binding and [^18^F]AV-1451 in these subcortical regions, although inconsistent findings are reported in studies using [^18^F]AV-1451 in PSP^8–12^. The sometimes lack of significant correlates of [^18^F]AV-1451 in PSP is often tribute to the low affinity of the ligand for 4R tau pathology, but in relatively small studies, the sessional variance of clinical rating scales may also reduce power. Therefore, our estimate of baseline clinical severity used the intercept extracted from the linear mixed effects model of longitudinal PSP-RS scores rather than single baseline assessment.

The null result for structural MRI predictors might be surprising given previous reports on the utility of visual and volumetric assessments of atrophy in midbrain and other subcortical regions, including caudate nucleus, putamen, globus pallidus, subthalamus and thalamus, as *in vivo* biomarker in patients with PSP^31^. Indeed, structural MRI has provided the most studied and validated diagnostic biomarkers in PSP. However, a biomarker’s properties for diagnostics (ie. presence of PSP^31,32^) or correlates of severity (i.e., at baseline) do not imply the property of prognostication.

Overall, our findings on the *in vivo* association between imaging markers of different pathological processes and their prognostic relevance accord with post-mortem data^2,3^, and suggest a key role for early microglial activation and tau burden on neurodegeneration, and consequent clinical progression. A growing literature supports a role for neuroinflammation in driving tau spreading and neurodegeneration in tauopathies^33–35^. Furthermore, genome wide association studies implicate inflammatory pathways in the etiology of tauopathies^36,37^. For example, Jabbari et al. reported an association between a common variation at the leucine-rich repeat kinase 2 (LRRK2) locus and survival from symptom onset to death in patients with PSP^37^. This relationship may be mediated by the effect of increased LRRK2 expression on microglia proinflammatory responses^38^, promoting spread and accumulation of misfolded tau protein, analogous to Alzheimer’s disease^35,39^. This hypothesis is supported by the association between dysregulation in expression of the microglial-related gene CXCR4, regional accumulation of neurofibrillary tangles and increased risk of PSP^40^. The role of early neuroinflammation in tauopathies is supported by PET evidence that microglial activation is observed before the PET evidence of aggregated tau and symptoms in carriers of MAPT mutations^41,42^. A preliminary study of longitudinal changes in microglial activation in two PSP patients showed stable microglial activation across a 6-10 months^4^, but may have lacked power.

There are several limitations to our study. We recruited according to clinical diagnostic criteria, and although clinicopathological correlations of PSP-Richardson’s syndrome are very high, including 8 of 8 cases in our study with *post mortem* pathology, they are not perfect. Moreover, the average rate of change in severity was 6-7 points per year on the PSP-RS, which is lower than several previous observational studies^20,43^ and clinical trials^44–47^. This difference may be partially explained by the selection criteria, favouring patients robust enough to undergo three brain scans at baseline. However, our cases were otherwise typical, and 15 out of 17 died within 5 years from baseline (mean 2.2 years ± 1.2). The modest size of our cohort prevented the application of complex models for the direct comparison between MRI and PET predictors, such as multiple linear regression or linear mixed models with several independent variables. The replication of these findings with larger and multicenter clinical cohorts will be an important next step to establish the generalizability of our results. Other limitations relate to the PET tracers used. [^11^C]PK11195 binds to the 18-kDa translocator protein which is overexpressed in activated microglia but also in other cell types, like astrocytes and vascular smooth muscle cells^48^, although [^11^C]PK11195 has been found selective for activated microglia over reactive astrocytes^49^. There are also caveats for [^18^F]AV-1451, with its off-target binding (monoamine oxidase, choroid plexus, neuromelanin) and lower affinity for PSP tau compared with Alzheimer related tau. Nonetheless, the topological distribution of [^18^F]-AV1451 binding and correlations with severity maintain utility for this ligand even in PSP^6,8–12,14^.

In conclusion, our results support the relevance of neuroinflammation to progression of PSP-Richardson’s syndrome. We suggest that [^11^C] PK11195 may be a valuable biomarker for clinical trials in PSP, complementary to structural MRI. The PET markers may be useful for stratification of patients based on prognosis, and the evaluation of therapeutic response, supporting the development immunomodulatory strategies for disease-modifying treatments in PSP alone or in conjunction with treatments directed against tau and other pathogenic pathways.

## Acknowledgments

We thank our participant volunteers for their participation in this study, and the radiographers/technologists at the Wolfson Brain Imaging Centre and Addenbrooke’s PET/CT Unit, and the research nurses of the Cambridge Centre for Parkinson-plus for their invaluable support in data acquisition. We thank the East Anglia Dementias and Neurodegenerative Diseases Research Network (DeNDRoN) for help with subject recruitment, and Drs Istvan Boros, Joong-Hyun Chun, and other WBIC RPU staff for the manufacture of the radioligands. We thank Avid (Lilly) for supplying the precursor for the production of [^18^F]AV-1451 used in this study. We also thank Frank Hezemans for consultation on statistical methods.

